# Spatial and single-cell transcriptomics landscape of adenomyosis

**DOI:** 10.1101/2025.07.06.25330330

**Authors:** Xing Yang, Chunjie Li, Junxian He, Bingbing Xie, Linyan Lv, Jian Xu, Yuan Lin, Chuanchuan Zhou, Manchao Li, Zhi Zeng, Jinfeng Tan, Shuqin Chen, Guizhong Cui, Guihua Liu, Shengbao Suo, Guangdun Peng, Xiaoyan Liang

## Abstract

Adenomyosis is a prevalent and non-cancerous uterine disease which can significantly impaired the fertility of reproductive-age women. However, the etiology, as well as the cellular and molecular mechanism underlying adenomyosis remain largely unknown. Here, we utilized cutting-edge spatial and single-cell RNA sequencing technologies to create a comprehensive transcriptional atlas of adenomyosis pathology. Our spatial profiling clearly distinguished gland, mesenchyma and myometrium regions, recapitulating spatial transcriptome structural characteristics of uterus. Moreover, we analyzed the expression profiles of 69,115 single cells and integrated them with spatial data. The analysis of immune cells showed a distinct immune inflammatory microenvironment in the eutopic and ectopic endometrial glands of adenomyosis. Notably, we discovered an increased number of *DNAH9*^+^ ciliated cells in ectopic endometrial glands, indicating their potential role in the formation of ectopic endometrium. These findings provide cellular evidence to support the invagination theory and offer a new vision on the pathophysiology and clinical intervention of adenomyosis.

## Introduction

Adenomyosis is characterized by the presence of endometrial glands and stroma-like tissue within the myometrium^1^, resulting in abnormal uterine bleeding, dysmenorrhea, chronic pelvic pain and infertility^2^. Around 35% of reproductive-age women suffer from adenomyosis worldwide^3, 4^. Highly variable symptoms make diagnosis challenging and commonly resulting in misdiagnosis. Furthermore, due to the advance of diagnostic imaging techniques such as transvaginal ultrasound scan (TVUS) and magnetic resonance imaging (MRI), there is an increasing trend of incidence rate, especially the remarkably increased proportion of young women diagnosed^4^.

Although several hypotheses have been proposed to explain the pathogenesis of adenomyosis, including invagination of the endometrium into the myometrium, trauma to the endometrial-myometrial interface, and the stem cell potential of de novo ectopic tissue^5^, none of the these theories can fully account for all the phenotypes^6^. This ambiguous understanding of underlying pathophysiologic mechanisms hinders the development of effective early intervention strategies, emphasizing the need for further research into the biological mechanisms involved.

Common symptoms of adenomyosis, such as dysmenorrhea and irregular uterine bleeding, suggest dysfunction of the endometrium, despite the absence of significant morphological change between in situ endometrium (eutopic) and the ectopic endometrial gland found within adenomyoma. Recently studies utilizing single-cell RNA sequencing (scRNA-seq) have reported functional differences within limited partial regions, offering some insight into cellular heterogeneity^7, 8^. It is important to note that these studies have been limited in scope, either focusing on a small number of patients or lacking investigation into the spatial distribution of specific disease regions. As a result, our understanding of adenomyosis at a high-resolution level remains incomplete. Advancements in scRNA-seq and spatial transcriptomics techniques provide powerful tools to investigate rare subpopulations, cellular interactions and tissue architecture^9–12^. These cutting-edge technologies have the potential to enhance our understanding of adenomyosis.

Herein, we simultaneously combined scRNA-seq and spatial transcriptome (Geo-seq)^13^ data to gain a global understanding of cell types and niches in the entire layer of adenomyosis uterus tissue. By allocating cell types to different adenomyosis niches, determining molecular mediators of intercellular interactions, and pinpointing the cellular and spatial sources of niche factors, our data shed new light on the invagination of eutopic endometrium and tissue architecture of adenomyosis. This multi-dimensional molecular profiling provides a foundation for further exploration of adenomyosis pathophysiology, enabling a better understanding of the cellular and molecular mechanisms involved. Ultimately, this study has the potential to facilitate early clinical diagnosis and medication intervention for adenomyosis.

## Results

### scRNA-seq and Geo-seq analysis of human endometrium and adenomyosis samples

scRNA-seq was performed on endometrial biopsies with the 10x Chromium system from eight individuals, including three individuals without adenomyosis and five individuals with adenomyosis (**Fig. 1a and 1b**). After quality control, we obtained a total of 69, 115 high-quality single cells by scRNA-seq, with a median 31,074 unique transcripts and 3,068 genes per cell **(Extended Data Fig. 1a)**. Based on their expression of known markers and the cell-type prediction with published models, we identified six main cellular categories: epithelial, endothelial, smooth muscle cells (SMC), fibroblast, myeloid cells, and lymphocyte (**Fig. 1c, 1d and Extended Data Fig. 1c**). These clusters were further grouped into 46 subclusters, highlighting the cellular complexity of both the endometrium and adenomyosis (**Extended Data Fig. 1b**). Overall, no significant difference was observed in cell-type composition between control and adenomyosis samples (**Extended Data Fig. 1d**), suggesting no newly discovered cell type in adenomyosis but mainly structural or functional alteration.

**Figure 1.**
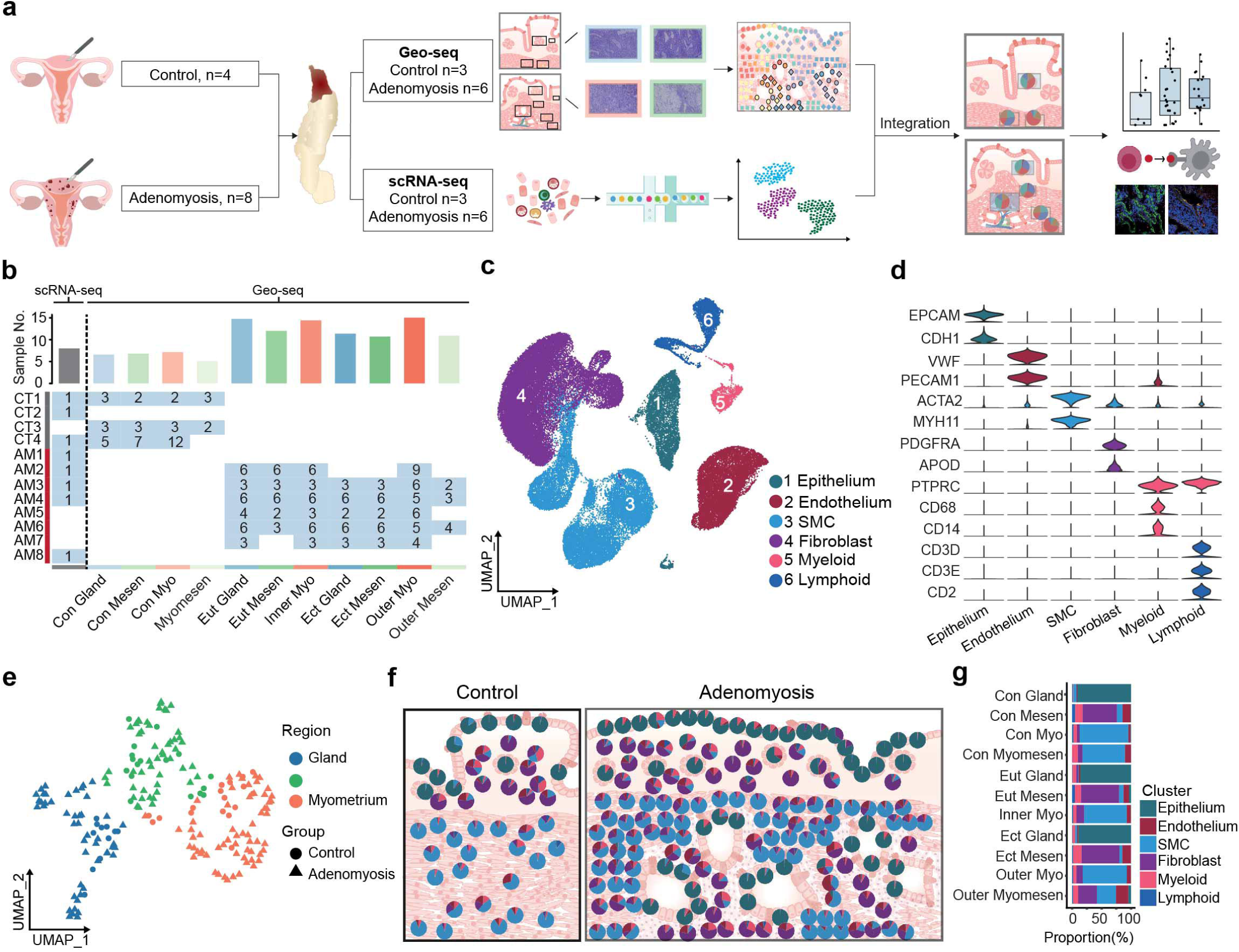

Since the ectopic endometrium of adenomyosis has different spatial information from eutopic endometrium, spatial transcriptome is important to further exploration of the pathophysiology of adenomyosis. To systematically dissect the tissue structure of endometrium, we performed laser microdissection based Geo-seq on 204 specimens collected from uterine tissues of nine individuals, six of whom had adenomyosis and three controls. We collected multiple spatial regions from control endometrial gland, control endometrial mesenchyma, control myometrium, control outer myometrial mesenchyma in control groups, and adenomyosis eutopic endometrial gland, eutopic endometrial mesenchyma, adenomyosis inner myometrium, ectopic endometrial gland, ectopic endometrial mesenchyma, adenomyosis outer myometrium, adenomyosis outer myometrial mesenchyma in adenomyosis groups, using precise morphology information to guide our sampling. (**Fig. 1b and Extended Data Fig. 2a** and 2b). Mapping of the raw reads onto human genome revealed that an average of around 15, 000 unique genes were detected in both the control and adenomyosis groups, and the samples exhibited similar expression density distribution and indicating high sample quality (**Extended Data Fig. 2c and 2d**). Uniform manifold approximation and projection (UMAP) of samples based on Geo-seq profiles clearly delineated gland, mesenchyma and myometrium regions, suggesting that the spatial transcriptome recapitulates the anatomical features of uterine lining (**Fig. 1e**).

To maximize the power of both techniques and reveal adenomyosis-associated cellular states that each technique individually may not detect, we integrated the scRNA-seq with the Geo-seq by deconvolution analysis based on the reference signature matrix produced by scRNA-seq with Cibersort algorithm^14^. As expected, gland regions were enriched with the epithelial cells, myometrium regions with SMCs, and mesenchyma regions with fibroblasts, endothelial cells, lymphocytes and myeloid cells (**Fig. 1f and 1g**). Together, these data indicated that tissue-structure associated cell types inferred by scRNA-seq and Geo-seq provided a proxy for dissecting molecular changes of tissue architecture and niches of adenomyosis.

### Characterization of ciliated cell in the ectopic endometrial gland of adenomyosis

Adenomyosis is characterized by the presence of endometrial glands within the myometrium of the uterus, with the main cell type of gland being epithelium (**Fig. 1g**). To gain insight into the molecular changes associated with adenomyosis, we performed further analysis of epithelium component. Based on the scRNA-seq data, we identified eight epithelial cellular subtypes based on the analysis of assigning their respective marker genes **(Fig. 2a and 2b)**. All of these clusters have been previously observed in endometrium^15, 16^, with the exception of luminal epithelium, which was absent given the low expression of known markers *SCGB2A2* and *WNT7A* (**Fig. 2b**). By integrating scRNA-seq and Geo-seq data, we defined two spatially distinct subsets: (1) *SOX9*^+^ epithelium enriched in both eutopic and ectopic endometrial glands of adenomyosis; and (2) ciliated epithelium mainly located in the ectopic endometrial glands of adenomyosis (**Fig. 2c**).

**Figure 2.**
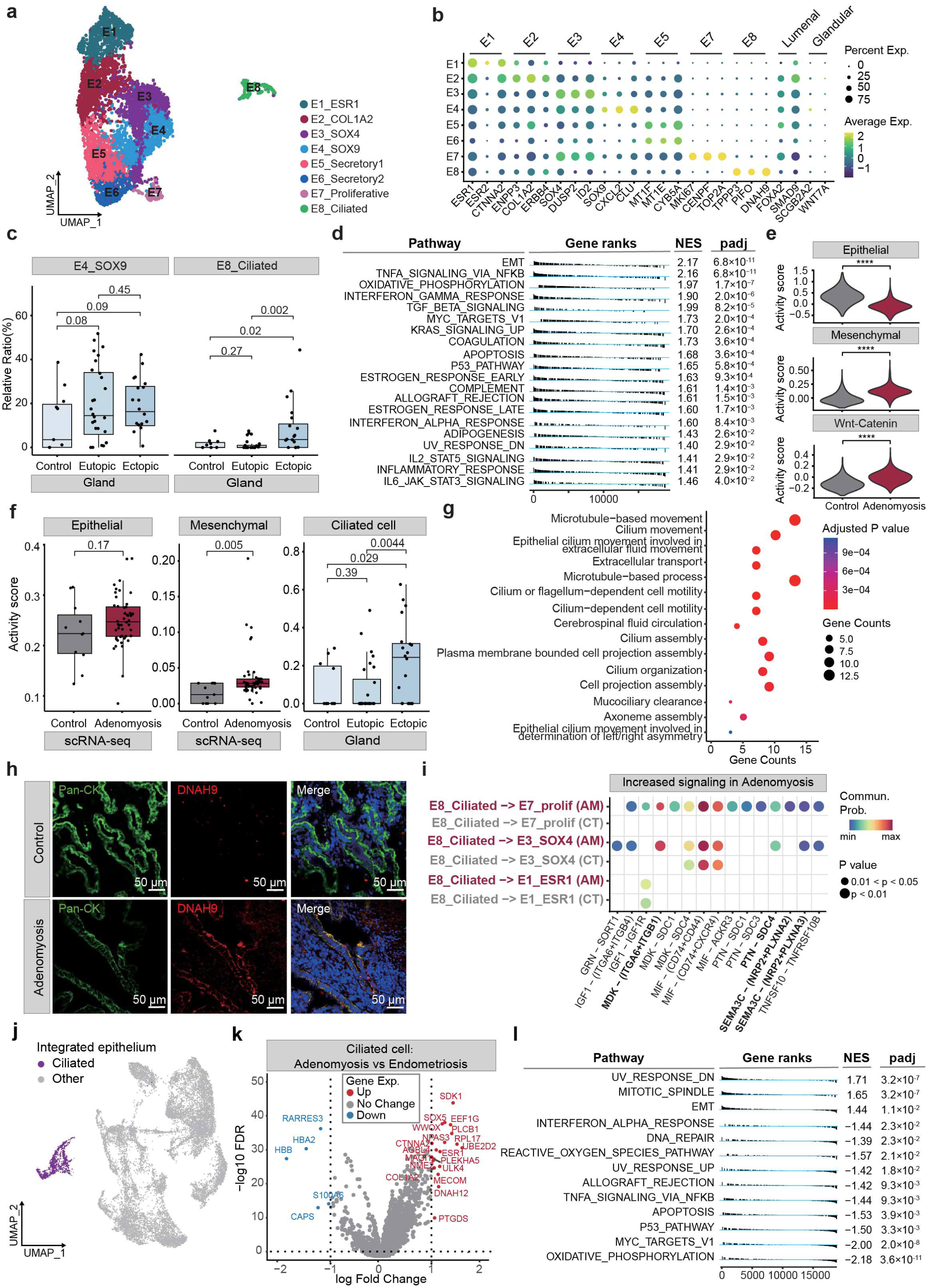

To further explore the role of *SOX9*^+^ epithelium in adenomyosis, we performed functional gene enrichment analysis. The top 15 expressed gene transcripts of *SOX9*^+^ epithelial clusters were significantly enriched in several biological processes, including inflammatory response and cell population proliferation. Additionally, pathways related to IL-17 signaling, NF-kappa B signaling and TNF signaling were prominently represented (**Extended Data Fig. 3a**), indicating extensively implicated inflammatory pathways in both eutopic and ectopic endometrial glands facilitate the symptomatology and the pathogenesis of adenomyosis.

To understand the biological characteristics between the endometrial glands from patients with or without adenomyosis, we conducted differentially expressed gene (DEG) and gene set enrichment analysis (GSEA) analysis based on Geo-seq data (**Extended Data Fig. 3b and Fig. 2d**). Of note, the samples from adenomyosis patients were significantly enriched in signatures related to epithelial-mesenchymal transition (EMT), TNF signaling pathway, interferon gamma response, TGF betta signaling, MYC targets V1, KRAS pathway, and apoptosis, which confirmed the inflammatory features in the glands of adenomyosis. Additionally, all of these pathways are related to estrogen signlaing^17^, suggesting hyperreaction of both eutopic and ectopic endometrium in adenomyosis patients in response to estrogen, and providing a new perspective of estrogen-dependent inflammatory schema. Among these pathways, EMT is critical in the pathogenesis of adenomyosis^18^. In the invagination theory, ectopic lesions originate from eutopic endometrium promoted by EMT. To confirm this, we quantified the epithelial and mesenchymal specificity scores in scRNA-seq (**Fig. 2e**) and in Geo-seq data (**Fig. 2f**). We found that the epithelium of adenomyosis present a higher mesenchymal score and lower epithelial score, and genes related to mesenchymal pathway were upregulated in adenomyosis (**Extended Data Fig. 3c**). In line with this, the WNT pathway, which was essential in EMT process, was highly activated in the adenomyosis group (**Fig. 2e**).

Intriguingly, our data revealed regional differences in the distribution of ciliated epithelium, suggesting potential functional differences between eutopic and ectopic endometrial glands. In the integrated data of the scRNA-seq and the Geo-seq, we used marker genes of ciliated cells (*TPPP3*, *PIFO*, *DNAH9*) as input regulons and found that the epithelium of ectopic endometrial gland scores significantly higher (**Fig. 2f**). We compared eutopic and ectopic endometrial glands based on Geo-seq data and found that samples of ectopic endometrial glands were enriched in epithelial cilium movement, cilium assembly and cilium organization (**Fig. 2g and Extended Data Fig. 3d**). Alternatively, mapping single cells to the spatial coordinates in tissue sections based on scRNA-seq and Geo-seq data by cellTrek algorithm^19^**(Extended Data Fig. 3h)**, we observed that ciliated cells were predominantly located in ectopic endometrial gland (**Extended Data Fig. 3f**). To investigate the potential role of ciliated epithelial cells, we applied the SColoc algorithm and identified spatial colocalization of ciliated cells with E7_proliferative, E3_SOX4 and E1_ESR1 (**Extended Data Fig. 3g**). We also identified the increased signal of ligand-receptor pairs between *MDK-ITGA6/ITGB1*, *PTN-SDC4* and *SEMA3C-NRP2/PLXNA3* in adenomyosis using cell-cell communication analysis (**Fig. 2i**). *SEMA3C*, a secreted glycoprotein, has been shown to drive EMT, invasiveness and stem-like characteristics in various types of tumors^20–23^. Finally, we confirmed the ciliated cells in ectopic endometrium through immunofluorescence (**Fig. 2h and Extended Data Fig. 3e**).

Furthermore, we performed a thorough comparison between our adenomyosis data and previously published scRNA-seq data on endometriosis. This published data also demonstrated a remarkable enrichment of signatures associated with ciliated epithelial cells^24^, which served as an additional motivation for us to delve into the functionality of these ciliated cells. By integrating the datasets, we were able to distinguish ciliated cells from other epithelial cells in both adenomyosis and endometriosis samples **(Fig. 2j and Extended Data Fig. 4a-b)**. Within these ciliated cells, we identified the specific expression of canonical marker genes *FOXJ1*, *PIFO* and *TPPP3*, further validating their identity (**Extended Data Fig. 4c**). Upon closer examination of gene profiles, we discovered distinct differences between the ciliated cells in adenomyosis and endometriosis. In adenomyosis, the ciliated cells exhibited a higher enrichment of gene programs associated with the EMT pathway, suggesting activation of the EMT pathway in ciliated cells potential involvement in the cellular process of invagination of endometrium **(Fig. 2j-l)**. Conversely, compared to endometriosis, the ciliated cells in adenomyosis demonstrated a lower activation of the interferon-alpha (IFNα) pathway, indicating a differential regulation of immune-related mechanisms **(Extended Data Fig. 4e)**. These results shed light on the unique characteristics and potential functional roles of ciliated cells in adenomyosis compared to endometriosis. Taken together, our findings revealed an ectopic gland-specific ciliated cell-type and profiled its cell signaling feature in adenomyosis. The ciliated cells in ectopic glands express a gene program specifying invasion and migration potential, consistent with the invagination theory of adenomyosis pathology. This process may drive EMT and multiple crosstalk similar to malignant tumors. Hence, targeting ciliated cell generation to inhibit the formation of ectopic lesions may be a novel potential therapeutic strategy.

### An inflammatory state in immune microenvironments of adenomyosis

Numerous studies have reported an aberrant immune microenvironment in adenomyosis^25, 26^. However, our scRNA-seq data only revealed a few changes in immune cells **(Extended Data Fig. 5a)**. Integration with scRNA-seq and Geo-seq data, on the other hand, showed significant alterations in lymphocyte and myeloid in the endometrial gland and mesenchymal regions (**Extended Data Fig. 2e**), highlighting the importance of spatial information in accurately delineating the microenvironment. We further explored the immune infiltration particulars in adenomyosis. scRNA-seq uncovered 10 lymphocyte subpopulations and 8 myeloid subpopulations, which were grouped into 18 cellular categories (**Fig. 3a and 3b**). The fraction of *CD8^+^* T cells in total immune populations was significantly increased, indicating a robust immune response (**Fig. 3c)**. Notably, elevated *CD8^+^* T cells in endometriosis tissue have been associated with an increased risk of infertility^27^.

**Figure 3.**
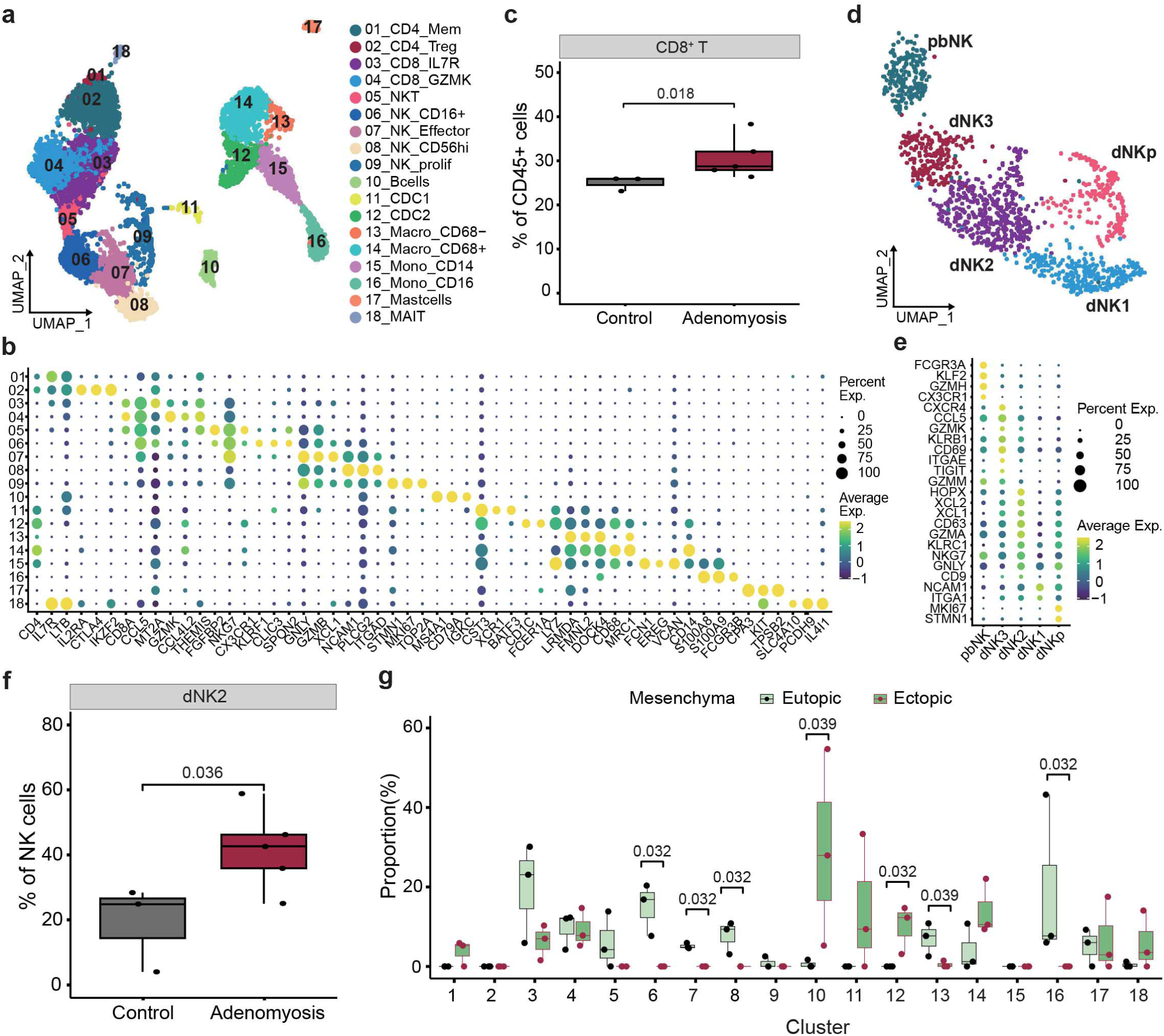

Natural killer (NK) cells play an essential role in fetal tolerance of uterine immune system^28^, but it remains elusive how the NK cells respond in adenomyosis. We identified five subclusters within NK cell population: three decidual NK (dNK1-3), one proliferation dNK (dNKp) and one peripheral blood NK (pbNK) (**Fig. 3d and 3e**). After calculating the ratio of each NK cluster in the patients, we found a significant enrichment of decidual dNK2 in adenomyosis samples (**Fig. 3f**). dNK2 cells expressed high levels of *XCL1* and *XCL2*, which encode two chemokines known to recruit *XCR1*^+^ cross-presenting dendritic cells (DCs) into tissues. These clusters also exhibited high level of *NKG7*, *GNLY*, *HOPX* and *CD63*, indicative of strong cytotoxic activity (**Fig. 3e**). With result of cellTrek **(Extended Data Fig. 3h),** we further identified an enrichment of NK_CD16^+^ and NK_effector in eutopic endometrium **(Fig. 3h)**, hinting that enrichment of cytotoxic NK cells in eutopic endometrium may be related to the high risk of infertility and miscarriage in adenomyosis patients. On the other hand, deficient of cytotoxic NK cells in ectopic endometrium suggested that weak immune surveillance is permissive for the survival of ectopic lesion in the myometrium.

Finally, we compared the differences of gene programs between control and adenomyosis samples in *CD8^+^* T cells and NK cells, respectively, but it did not reveal significant differences in gene profiles between samples from control and adenomyosis samples (**Extended Data Fig. 5b and 5e**), suggesting that the *CD8^+^* T cells and NK cell-specific gene programs do not change significantly in response to the disorder, but rather the populations themselves shift in frequency. Other *CD4^+^* and *CD8^+^* T cells showed no regional heterogeneity among control, eutopic and ectopic mesenchyma as well (**Extended Data Fig. 5c)**.

Macrophages and DCs have been described as key players in endometriosis pathology^29^, and we sought to evaluate the cellular mechanisms of macrophage and DCs in adenomyosis. In the integrative data, we identified an enrichment of conventional type 1 dendritic cells (cDC1) in both eutopic and ectopic regions of adenomyosis tissue (**Fig. 4a**). cDC1 is crucial for antitumor immunity and their abundance within tumors is linked to immune-mediated rejection and the success of immunotherapy^30^. *XCR1*, a chemokine receptor expressed by cDC1, is recruited by *XCL1*^+^ dNK2 cells. The expression pattern of the *XCL1/2*-*XCR1* ligand-receptor pair further confirmed functional interactions between dNK2 and cDC1 in our analysis (**Fig. 4b**). DEGs analysis also revealed that cDC1 highly expressed genes related to inflammatory responses, such as *BIRC3*, *TNFAIP3*, *IL1R2*, *JUN* and *IDO1* (**Fig 4c**). An enrichment of *CD68^+^* macrophages in adenomyosis samples was also detected (**Fig. 4d**). To further understand the characteristics of these macrophages, we scored the expression of genes related to M1/M2 phenotypes, proliferation, phagocytosis and angiogenesis^31^. The result revealed that *CD68^+^*macrophages were more similar to M2 phenotype and exhibited strong phagocytotic activity score (**Fig. 4e**). Additionally, we observed that macrophages from adenomyosis exhibited stronger phagocytosis and weaker angiogenesis, although the difference was not significant (**Fig. 4f**). Together, these findings suggested that adenomyosis is associated with an inflammatory immune microenvironment enriched with *CD8^+^* T cells, dNK2 and showed heterogeneity of regionalized of cytotoxic NK cells, cDC1, and *CD68^+^* macrophages, which indicates intense phagocytose and immune cytotoxic reaction to reject ectopic endometrial islands and potential cellular mechanism of infertility in patients with adenomyosis.

**Figure 4.**
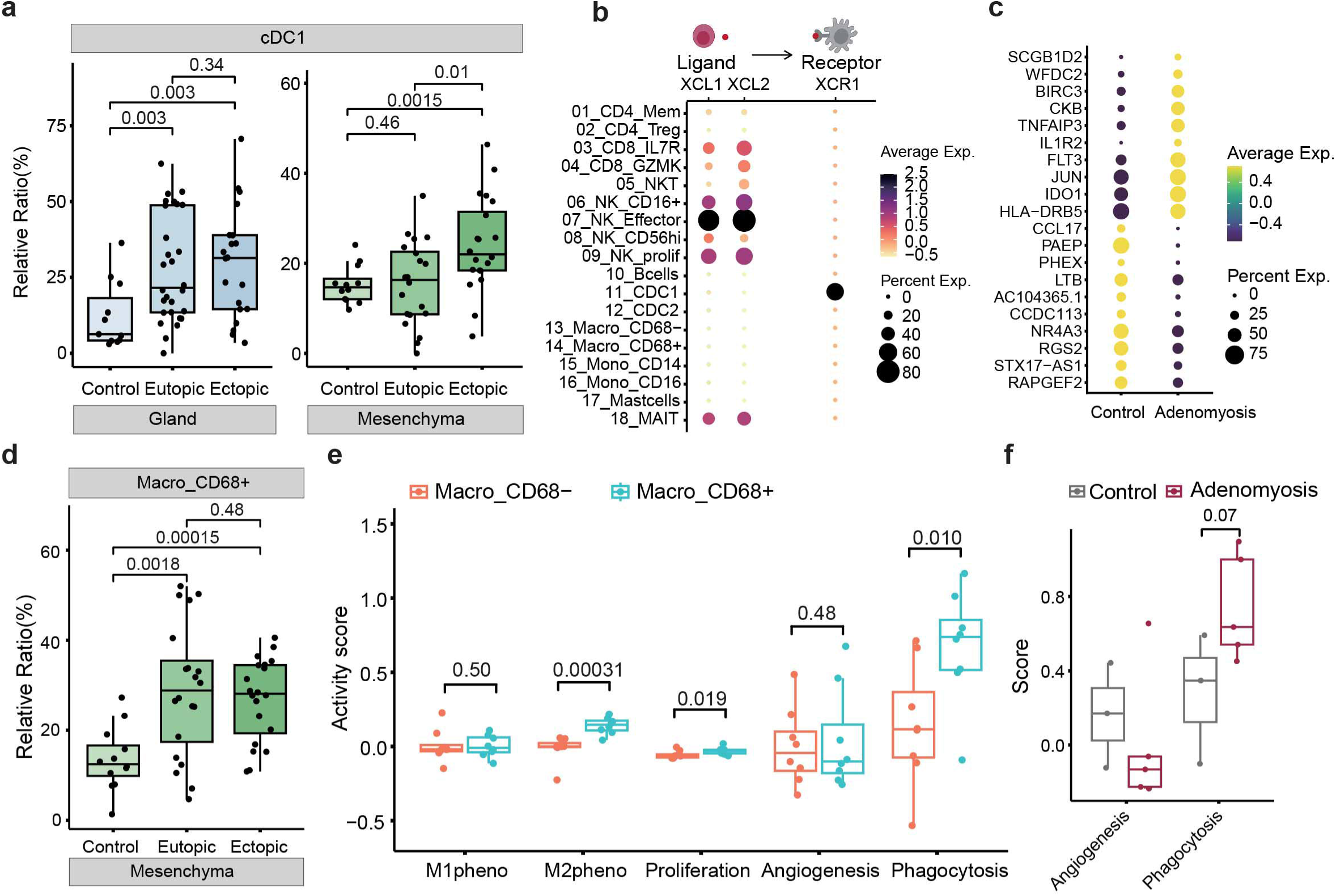

### Similarity between cancer-associated and adenomyosis-associated fibroblasts

Adenomyosis is characterized by the overgrowth of fibrous tissue^32^ and the process of endometrial basalis invasion is similar to the metastasis of malignant tumors. Also, we found that fibroblasts increased significant in adenomyosis inner myometrium and outer myometrium (**Extended Data Fig. 2e**). To understand the diversity of fibroblast in adenomyosis, we examined their heterogeneity and identified six populations (**Fig. 5a**). Among these populations, three were decidualized fibroblast clusters (dFibro-S100A4, dFribo-PLCL1, and dFibro-NKB1) expressing *CFD*, *FOXO1* and *IL15*, while the remaining two were endometrial fibroblast clusters (eFibro_MMP11 and eFibro_prolif) expressing *MMP11*, *CRABP2* and *ECM1* (**Fig. 5b**). We compared the deconvoluted proportions of each cluster in mesenchymal regions and found that the enrichment of eFibro_MMP11 and eFibro_prolif populations was significantly higher in both eutopic and ectopic mesenchyma of adenomyosis than in control samples (**Fig. 5c).** A previous study has shown that *MMP2* and *MMP9* expression may play a critical role in adenomyosis development by facilitating the invasion of endometrial tissues into the myometrium and promoting angiogenesis in adenomyotic implants^33^. Additionally, *MMP11* has been found to promote the proliferation and progression of various types of tumors^34, 35^. The eFibro_MMP11 population also highly expressed *TNC*, a marker protein for cancer-associated fibroblasts (CAFs) (**Fig. 5b**). CAFs play a crucial role in cancer pathogenesis by remodeling the extracellular matrix structure and facilitating the invasion of the tumor cells^36^. Pathway analysis revealed the enrichment of focal adhesion, ECM-receptor interaction, PI3K-AKT signaling pathway and gap junction based on the marker genes of eFibro_MMP11 cluster (**Fig. 5d**). On the other hand, the eFibro_prolif population was characterized by co-expression of proliferation markers. Of note, we found a decrease of cell proportion in the dFibro-S100A4 cluster in both eutopic and ectopic mesenchyma of adenomyosis (**Fig. 5c**). Successful pregnancy requires fibroblast differentiation into specialized decidual cells^37^. Dysregulation of decidualized fibroblast and an increase in proliferation fibroblast may shed light on the potential cellular mechanism of infertility in patients with adenomyosis.

**Figure 5.**
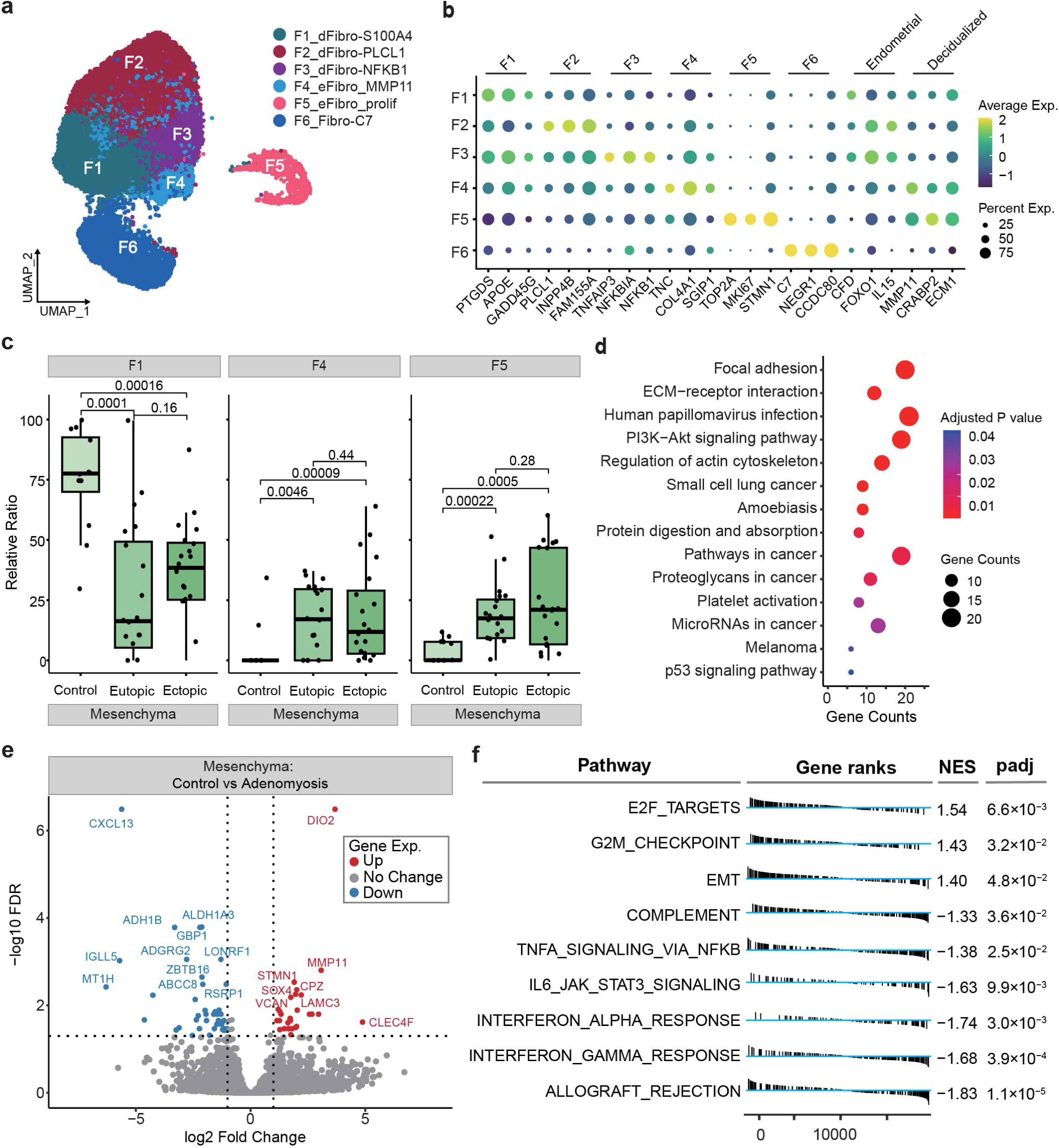

Subsequently, we compared the gene profiles between eutopic and ectopic endometrial mesenchyma using Geo-seq data. The results indicated highly expressed genes related to angiogenesis such as *MMP11*, *LAMC3*, *SOX4*, *VCAN*^38–43^ and the enrichment of E2F targets, G2M checkpoint and EMT pathway in adenomyosis samples (**Fig. 5e and 5f**). This is consistent with our deconvolution result, which showed a high proliferation status in fibroblast from adenomyosis, resulting in increased stiffness and thickness in the uterine tissue. Together, our data suggest the presence of CAFs and proliferation gene signatures in fibroblasts of adenomyosis may facilitate the invagination of the endometrial basalis process. Therefore, targeted treatment to reduce fibrogenesis may represent a promising intervention strategy for adenomyosis.

### Role of SMCs diversity in angiogenesis in adenomyosis

Adenomyosis is a condition characterized by the presence of islands of endometrial tissue surrounded by hypertrophic smooth muscle cells (SMCs) within the myometrium. To better understand the heterogeneity of SMCs, we divided them into nine subclusters (**Fig. 6a** and **6b**). Functional enrichment analysis showed that SMCs in adenomyosis was enriched for EMT and apical junction signatures compared to control samples (**Fig. 6c**). Furthermore, we observed an enrichment of vascular smooth muscle cell populations (S01_vSM_1 and S02_vSM_2) in myometrium of adenomyosis (**Fig. 6d**), suggesting an alternative microenvironment with abundant blood vessels in adenomyosis. Perivascular *MYH11* (Prv-MYH11) cells, previously characterized in myometrium^44^, were found to be enriched in control myometrium. We also performed DEGs and GO analysis between control and myometrium samples from Geo-seq data, revealing highly expressed genes related to angiogenesis such as *SOX4*, *ASPN*, *MMP10*, *F3*^42, 45–48^ and enriched GO terms related to the WNT pathway and cellular response to growth factor stimulus (**Fig 6e and 6f**). Together, these data suggested that angiogenesis is a prominent feature of adenomyosis. The marked increase in angiogenesis within the adenomyosis may further complicate the disease by creating a favorable environment for gland proliferation and invasion, which exacerbates the symptoms and causes deleterious effects of the adenomyosis. Consequently, intervention in the angiogenic process represents a critical therapeutic avenue for the management of adenomyosis.

**Figure 6.**
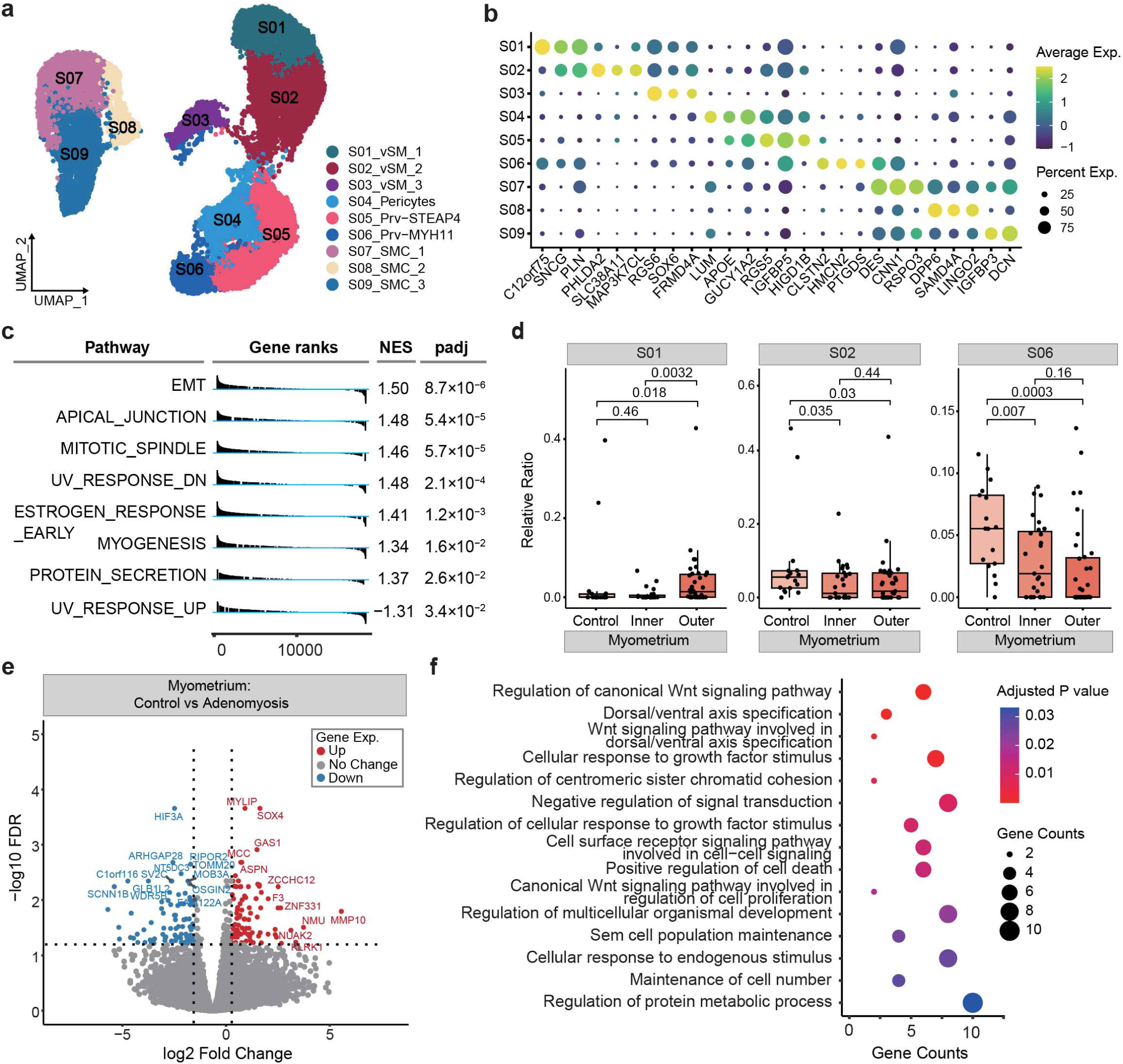

## Discussion

Studies on the pathogenesis of adenomyosis are constantly evolving, but due to the limitations in procuring clinical specimens and the difficulty in modeling it *in vitro*, the exact mechanisms remain mystery. Previous studies on the endometrium of women with adenomyosis have typically focused on the expression of single gene or a limited number of genes^6, 49^. Recently, Sule et al. have performed single-cell analysis and illustrated profound cellular heterogeneity in adenomyosis^8^. But they only selected control, eutopic endometrium and adenomyotic tissues, which is unable to reveal the landscape changes of adenomyosis. To address this gap in knowledge, our study employed scRNA-seq of the entire layer of uterine samples to conduct a more comprehensive transcriptional profiling of adenomyosis compared to previous studies^7^. In addition, we retained the spatial information of tissue organization by profiling the transcriptome in representative regions of adenomyosis through low-input spatial RNA-seq analysis. By mapping scRNA-seq data onto spatial transcriptome with deconvolution, we explored the genome-wide transcriptome at single-cell resolution. Several cell subclusters involved in the formation of ectopic endometrium were identified, therefore this study offers novel insights into the cellular mechanism of adenomyosis.

Epithelial cells play a crucial role in the function and pathology of endometrium. scRNA-seq data analysis of adenomyosis conducted by Liu *et al.*^7^ showed that endometrium in adenomyosis has an inflammation feature, which is consistent with our findings. Additionally, our integrated spatial information identified higher composition of *SOX9^+^*and ciliated epithelial cells in both eutopic and ectopic endometrial glands, expressing genes related to inflammation. *SOX9^+^* epithelial cells have been shown to play a role in the formation of endometrial cancer^50^ and ectopic endometriosis lesions ^51^. Previous scRNA-seq studies have also identified ciliated cells and showed distinct transcriptome profiles from other epithelial cells and high activity of WNT pathway targets^15, 24, 44^. Our study confirms the presence of ciliated cells in situ endometrium and ectopic endometrium, but with significantly higher proportions in ectopic endometrial glands than in eutopic and control endometrial glands, as confirmed by immunofluorescence. Moreover, the DEGs in the epithelial cells of adenomyosis are significantly enriched in the pathways and terms related to the movement and assembly of ciliated cells. The interaction between *SOX4^+^* epithelial cells, which known as a marker of high proliferation and EMT^52^, is increased in adenomyosis. Therefore, ciliated cells may play an important role in the proliferation of ectopic endometrium. However, the causal relationship between increased *SOX9^+^* and ciliated epithelial cells in adenomyosis remains unclear and awaits further in-depth investigation. Adenomyosis is a hormone-dependent disease and estrogen (E2) signaling has been shown to drive ciliogenesis in the endometrium in endometrial organoid^53^. We speculate that ciliated cells may proliferate in ectopic lesions driven by estrogen and promote the proliferation of ectopic endometrium.

The inflammation at the endo-myometrial interface may facilitate the invasion of endometrium into the myometrium, forming adenomyosis lesions^37^. Consistently, we found that in *SOX9^+^* epithelium clusters, highly expressed genes were enriched in inflammation and apoptotic signaling. Imbalanced inflammation and apoptotic signals have been reported in adenomyosis^25, 54–56^. The ectopic endometrium may activate the inflammation and apoptotic response and even lead to infertility in patients with adenomyosis. Together with the inflammatory status of immune microenvironment, we encision that treatments targeting inflammation and apoptotic may relieve symptoms for patients with adenomyosis. Additionally, our study has further verified adenomyosis to associate with an inflammatory immune microenvironment, which closely related to the development and progression of the disease. The immune system is thought to play a vital role in uterine cavity microenvironment. On the one hand, it is essential to protect against pathogen invasion by way of an appropriate inflammatory reaction, while on the other hand, a shift toward an immunosuppressive state is also required to allow successful embryonic implantation. NK cells, macrophages, T cells and dendritic cells are modulated by the uterine microenvironment, and in turn, contribute to tissue homeostasis^28, 57^. The inflammation feature of glands suggested that the active immune response in adenomyosis involves *CD8^+^* T cells, which is consistent with previous study, demonstrating that the proportion of *CD8^+^* T cells is increased in the eutopic endometrium of women with endometriosis^27^. Similarly, Liu applied flow cytometry to investigate the alterations in immune cell subsets in adenomyosis and found an increase of *CD8^+^* T cells was the predominant alteration in ectopic lesions in patients with adenomyosis and was significantly associated with the severity of adenomyosis^58^. All these suggest that therapeutic strategy designed to target *CD8^+^* T cells may be beneficial to adenomyosis patients.

As part of the lymphocytes, NK cells are a crucial component of the innate immune response and they can rapidly eliminate tumors or infected cells without targeting normal cells^59^. Uterine NK cells are the predominant leukocyte population in human endometrium, comprising 30-40% of the total leukocyte population in the proliferative phase^60^. As the most abundant immune cell population in the uterus, NK cells in uterine are specifically denoted as decidual NK cells (dNK). Evidence is accumulating that NK cells are involved in the pathogenesis of adenomyosis and endometriosis. Kanzaki *et al*. ^61^ showed that serum obtained from patients with endometriosis can inhibit NK cells activity *in vitro*, and it is reasonable to speculate that decreased NK cells activity may be associated with the onset or progression of endometriosis. Moreover, several studies have identified that ectopic endometrium may escape NK cell-mediated clearance, and the impairment of NK cell function may confer an immune-privileged status to the extra endometrium growth^62–65^. However, different types of NK subpopulation respond differently to stimulation. To elucidate the role of NK cells in adenomyosis, we further divided NK cells into pbNK, dNK1, dNK2, dNK3 and dNKp subpopulations based on previous markers^28^. Our data showed that the proportion of dNK2 is significantly higher in adenomyosis patients than in the control samples. dNK2 highly expresses inflammatory cytokine genes, such as *XCL1*, *XCL2* and *GZMA*. Interestingly, we found that cytotoxic NK cells are increased in eutopic endometrium while deficient in ectopic endometrium. Together, it is reasonable to speculate that abnormal growth of ectopic lesions can trigger the activation of *CD8^+^* T cells and dNK2. However, the presence of cytotoxic NK cells, found only in eutopic endometrium, may hinder successful pregnancy implantation and make it harder to remove the ectopic lesions.

Macrophages play key roles in both innate and adaptive immune responses as they are involved in phagocytosis, antigen presentation to T cells and the secretion of cytokines/chemokines. In normal endometrium, macrophages represent approximately 10% of the total endometrial immune cell population^66^. While recent study has proposed that macrophages and DCs present an immunotolerant phenotype in the endometriosis microenvironment^16^, little research has been conducted on the microenvironment of adenomyosis myometrium. Similar to previous study on endometriosis^16^, we also found that there were more cDC1 in eutopic and ectopic endometrium. Furthermore, the increased cDC1 may be recruited by dNK2 through XCL1/2-XCR1. Several lines of evidence suggest that macrophages are not only “trapped” in ectopic endometrial lesions but also locally activated^67, 68^. Lousse *et al.* confirmed that the nuclear factor-κB transcription factor is activated in macrophages from endometriotic patients, and responsive genes that determine macrophage functions undergo transactivation, supporting angiogenesis and tissue remodeling^69^. Monica et al. found that macrophages infiltrate into ectopic lesions, modulate adaptive immune response, promote the growth of extra endometrium and induce angiogenesis^70^. An et al used eutopic endometrial cells from adenomyosis-affected uteri to co-culture with macrophages and found that macrophages switched to M2 phenotype^25^. Consistent with the previous study^16^, we also found that the proportion of *CD68^+^* macrophages was higher in both eutopic and ectopic endometrium, exhibiting M2 phenotype feature. However, *CD68^+^* macrophages didn’t show the activation of angiogenesis, but revealed strong phagocytotic activity. Further studies are needed to determine the function of *CD68^+^* macrophages in adenomyosis.

Studies have suggested that adenomyotic lesions progress to fibrosis through various mechanisms, including epithelial-mesenchymal transition, fibroblast-to-myofibroblast transdifferentiation and smooth muscle metaplasia^32, 71, 72^. However, the role of fibroblast in adenomyosis remains poorly characterized. In our study, we provided a more precise characterization of fibroblast and identified a novel fibroblast subpopulation that may contribute to the invasion of endometrial tissues into the myometrium. We found that the eutopic and ectopic endometrial mesenchyma contained more eFibro_MMP11 cells, a subpopulation that shares the same marker gene with CAFs. CAFs are the most abundant stomal cells in cancer, and are known to modulate cancer progression and promote metastasis^73^. Enrichment analysis of DEGs in fibroblasts showed significant enrichment in focal adhesion and ECM-receptor interaction pathways, further implicating the role of fibroblasts in the invasion of endometrial tissues.

Adenomyosis is a pathology associated with sophisticated molecular mechanisms. We presented a comprehensive map of the cellular landscape of adenomyosis combined Geo-seq and scRNA-seq technologies, which serves as a valuable tool for dissecting the molecular changes in tissue architecture. As we proposed in **Extended Data Fig. 6**, adenomyosis can result from multiple pregnancies and iatrogenic factors, leading to the invasion of ciliated cells into the myometrium. The ectopic endometrium may escape NK cell-mediated clearance due to its immune-privileged status resulting from reduced NK cells, and thus survive in the uterine myometrium. However, this survival can activate the immune response, causing fibrosis and angiogenesis. Additionally, the cytotoxic NK cells in the eutopic endometrium may contribute to the infertility in patients with adenomyosis. Adenomyosis is not only a disease of abnormal endometrial invasion but also involves an imbalance of immune environment and the movement of ciliated cells. Our analysis therefore highlighted the potential of modulating immune environment and ciliated cells as therapeutic strategies, albeit in-depth investigation of regulatory mechanism in various modeling system required.

## Method

### Patient materials

This study was approved by the Ethics Committee of the Institutional Review Board at the Sixth Affiliated Hospital of Sun Yat-sen University and conducted according to all relevant ethical regulations regarding human participants. Written informed consents were obtained from all participants in the study.

### Surgical approach and tissue collection

Patients diagnosed with adenomyosis were treated with hysterectomy surgery. For the control, we included individuals without the diagnosis of adenomyosis, and underwent hysterectomy surgery. Adenomyosis was diagnosed based on hematoxylin and eosin (H&E) stained tissue slides. Complete clinical information for participants is provided in Supplementary Table 1. After confirmation of H&E slides, fresh tissue was stored in MACS tissue storage solution (Miltenyi, #130-100-008) and kept on ice until following processing.

### Tissue Laser Capture Microdissection and RNA isolation

Uterine tissue of adenomyosis and control patients were embedded in optimum cutting temperature (OCT) compound, snap-frozen in liquid nitrogen and labeled the direction. After the OCT was completely frozen, the tissues were cryosectioned at 20 μm. Geo-seq procedure was followed as described previously^74^. Control endometrial gland, control endometrial mesenchyma, control myometrium, control myometrial mesenchyma in control samples, and eutopic endometrial gland, eutopic endometrial mesenchyma, adenomyosis inner myometrium, ectopic endometrial gland, ectopic endometrial mesenchyma, adenomyosis outer myometrium, adenomyosis outer myometrial mesenchyma in adenomyosis samples were captured by laser microdissection and subjected to the BGI2000 sequencer with a 150-bp paired-end model.

### Immunofluorescence

Frozen sections were cryosectioned at 5 μm. Slides were incubated for 10 min at 60 °C in a dry oven. Tissue was permeabilized with 0.1% Triton X-100 in PBS for 10 min and blocked with 5% BSA for 1h at room temperature, then incubated with primary antibodies against DNAH9 (#11210R; Shanghai Yaji Biotechnology, Shanghai, China,1:100 dilution) and pan-cytokeratin (#ab7753, Abcam, 1:100 dilution) at 4°C overnight. Tissue sections were subsequently incubated with secondary antibody goat anti-mouse Alexa Fluor 488– (#A-31573,Invitrogen, 1:100 dilution) and Cy3-labeled goat anti-rabbit (#BA1032; Boster, Wuhan, China, 1:100 dilution) for 1 h at 37 °C. 4,6-Diamidino-2-phenylindole (DAPI) (#D9542, Sigma, 20 µg/ml dilution) was used to counterstain the nuclei, then the slides were mounted with Mounting Medium (Solarbio, S2100). Images were captured with a Zeiss Laser Scanning Microscopes.

### Tissue Preparation for single-cell RNA-seq

Fresh tissues were used for single-cell RNA-seq. Uterine tissues were divided into endometriosis and myometrium. They were separately dissociated into single cell in dissociation solution (0.35% collagenase IV5,2 mg/mL papain, 120 U/mL DNase I) in 37LJ water bath with shaking for 20 minutes at 100 rpm, and digestion was terminated with 1x PBS containing 10% fetal bovine serum (FBS, V/V). Then, the cell suspension was filtered by 70-30μm stacked cell strainer and centrifuged (300g,5 min,4LJ). Red blood cells were removed with Red Blood Cell Lysis Solution (MACS 130-094-183, 10x). Dead cells were removed with Miltenyi ® Dead Cell Removal Kit (MACS 130-090-101). Viable cells were counted with haemocytometer/ Countess II Automated Cell Counter and concentration adjusted to 700-1200 cells/μL.

### Single-cell capture, library preparation and sequencing

Single-cell suspensions were loaded to 10x Chromium to capture single cell according to the manufacturer’s instructions of 10x Genomics Chromium Single-Cell 3’ kit(V3). Libraries were sequenced on Illumia NovaSeq 6000 sequencer at a depth of 20,000 reads per cell.

### Single-cell data pre-processing and clustering

The Cell Ranger toolkit (v.7.0.0, 10x Genomics) was used to demultiplex the FASTQ reads align raw reads to the human reference genome (GRCh38, 10x Genomics), and generate the gene-cell unique molecular identifier (UMI) matrix for each sample. The resulting count matrices were further processed using Seurat packages (v.4.3.0) to exclude poor quality cells throuth the following steps. First, we filtered cells with high mitochondrial gene expression (>12.43%) by fitting the expression of mitochondrial genes to a normal distribution and applying a false discovery rate (FDR) threshold of <0.01. Second, we removed cells with a low number of genes detected, specifically those with fewer than 300 genes. This step helped eliminate low-quality cells that may have been subject to technical artifacts. Third, we employed the DoubleFinder (v. 2.0.3) to identify potential doublets and used a cutoff of the 92.5th percentile for the doublet score. Cells exceeding this threshold were considered potential doublets and were subsequently removed from the analysis. Following these quality control steps, we obtained a final dataset comprising 69, 115 single cells for the following analysis.

To integrate the individual samples and identify common sources of variation, we performed canonical correlation analysis (CCA). The data was normalized by using “NormalizeData” function, ensuring that the number of UMIs in each cell was equal to the median UMI across the entire dataset. Additionally, a log-transformed was applied to the data. “FindVariableFeatures” function was used to identify the top 2,000 highly variable genes. Subsequently, the gene expression matrix was scale and center using the “ScaleData” function. For dimensionality reduction and visualization of the data, we performed principal component analysis (PCA) using the RunPCA function based on the highly variable genes. The resulting top 30 principal components were then used for uniform manifold approximation and projection (UMAP) analysis to visualize the clusters in 2-dimensional space. Finally, clustering was performed using the Leiden community detection algorithm, which allowed us to identify distinct cell populations based on their transcriptional profiles.

### Annotation of cell clusters

To assign the major cell types for each cluster, we followed a two-step approach. First, we performed differential expression analysis using “FindAllMarkers” function with default parameters. By comparing each cluster against all other clusters, we identified genes that showed significant differential expression. In the next step, we aimed to assign cell types to the clusters by examining the presence of known cell-type-specific genes among the top rank of differentially expressed genes in each cluster. By referencing established knowledge about marker genes for specific cell types, we determined the most likely cell type for each cluster based on these differentially expressed genes.

For the annotation of sub-cluster of immune cells, we applied an unbiased cell type recognition method named SingleR (v.2.0.0), which leverages reference transcriptomic datasets of known cell types for annotation. We first applied SingleR to assess if the predicted annotations for a particular query cluster were consistent across different reference datasets. Based on this analysis we assigned the predicted cell type annotation to the cluster. Alternatively, we also checked the genes present in the top-rank list of differentially expressed gene for that cluster to aid in the annotation process.

### Geo-seq data analysis

The raw reads from Geo-seq underwent a series of pre-processing steps. First, a quality control check was performed using FastQC (v.0.11.9) to assess the quality of reads. Subsequently, adapter sequences were trimmed sequentially using Trim Galore (v.0.6.7). The clean reads were then aligned to the human reference genome (GRCh38) using Hisat2 (v.2.2.1). Samtools (v.1.17) was used to create indicesand merge BAM files of reads. For transcript expression estimation, Stringtie (v.2.1.7) was utilized. This tool leveraged the GTF annotation and bam alignment to estimate the expression levels of transcripts. The prepde.py3 script from Stringtie was used to extract counts information and to calculated transcripts per million (TPM) of each gene. The TPM value considers both the length of the gene and the read count mapped to that gene, providing a normalized measure of gene expression.

### Differential expression analysis for Geo-seq data

Differentially genes expression (DGE) between patients with and without adenomyosis was performed on theGeo-seq data using the popular packages edgeR (v. 3.40.2) with default parameters. DGE was defined as genes that exhibited an adjusted *p* < 0.05 and log2(fold change) > 1, unless specified otherwise.

### Functional enrichment analysis of gene set

To evaluate the adenomyosis associated gene programs, we employed gene set enrichment snalysis. Specifically, we utilized the “enrichGO” function from the clusterProfiler package (v. 4.6.2) to evaluate the fold enrichment score and p-value of differential genes within various Gene Ontology (GO) categories, including biological process (BP), cellular component (CC) and molecular function (GO-MF), as well as Kyoto Encyclopedia of Genes and Genomes (KEGG) pathways. This analysis allowed us to determine whether specific gene sets or pathways were significantly enriched among the differentially expressed genes.

In addition, we also performed the fast gene set enrichment analysis using fGSEA (v.1.11.1, R package). This analysis focused on identifying common pathways using the hallmark gene sets in the molecular signatures database (MsigDB). For all enrichment analysis, adjusted *p* < 0.05 was regarded as statistically significance, indicating the enrichment of specific gene sets or pathways in relation to adenomyosis.

### Calculation of gene set activity score

To assess the enrichment of a specific gene set or other relevant gene sets among the highly expressed genes in each cell, we employed a method called AUCell. AUCell^75^ is specifically designed for scRNA-seq data analysis. It allows for the identification of cells with active gene regulatory networks by quantifying the enrichment of a gene set within individual cells. The AUCell method calculates a gene set activity score for each gene set by measuring the area under the recovery curve (AUC) across the ranked expression values of top expressed genes (5% in default) within a given cell. This score reflects the proportion of expressed genes in the query gene set and their relative expression levels compared to other genes within the cell. Importantly, this approach is not influenced by the units of gene expression or the normalization method used, providing a robust and unbiased measure of gene set activity. We focused on evaluating the activity scores of gene sets related to epithelial mesenchymal and endothelial mesenchymal transitions, WNT pathway and ciliated cell markers (*TPPP3*, *PIFO* and *DNAH9*) in each cell.

### Deconvolution analysis of Geo-seq data using CIBERSORT

In order to compare the profile of patients with adenomyosis and controls in each Geo-seq sample, we utilized the CIBERSORT algorithm, which allows for the estimation of cell-type composition within each tissue location by Using Geo-seq transcriptomics data and scRNA-seq signature matrix. The scRNA-seq signature matrix (denoted as B) is generated using “FindAllMarker” function in Seurat pacakage and serves as a reference. It consists of K cell types, each characterized by a set of G cell-type-informative genes. In the Geo-seq transcriptomics data, we denote the G-by-N gene expression matrix as X, where G represents the same set of informative genes and N represents the number of spatial locations. We also denote the N-by-K cell-type composition matrix as V, where each row of V represents the proportions of the K cell types within each spatial location. To estimate V, the CIBERSORT algorithm employs a non-negative matrix factorization model to link the three matrices:

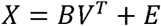

where E is a G-by-N residual error matrix.

CIBERSORT algorithm applied a novel machine learning method of nu-support vector regression (ν-SVR) to estimate matrix V, which outperformed other approaches^14^. This analysis provided valuable spatially insights into the cellular heterogeneity and potential differences in cell-type proportions between the two groups, contributing to our understanding of the cellular dynamics associated with adenomyosis.

### Spatial mapping of single cells using CellTrek

CellTrek^19^ is a powerful tool used to accurately map single cells back to their spatial coordinates within tissue sections, utilizing both scRNA-seq and Geo-seq data. This innovative approach involves several key steps outlined in the CellTrek tutorial. Briefly, the “traint” function was used to co-embed scRNA-seq and Geo-seq datasets. This process enables the integration of information from both datasets, facilitating the mapping of single cells to their spatial locations. Subsequently, the “celltrek” function was utilized with default parameters to chart single cells to their respective spatial locations. This step leverages the co-embedding information to accurately position each cell within the tissue section. We used the non-linear interpolation approach to augment the spatial spots, resulting in a more precise mapping of the single cells to their corresponding locations within the tissue. Finally, the obtained results were further analyzed using the SColoc module, which examines the colocalization patterns between different cell types. By applying default parameters, this module provides valuable insights into the spatial relationships and interactions between various cell types within the tissue.

### Ciliated epithelium integration analysis with endometriosis dataset

scRNA-seq data of endometriosis was downloaded from NCBI Gene Expression Omnibus (GEO) under accession number GSE213216. To assign cell types within this dataset, we utilized the markers *EPCAM* and *CDH1* to identify epithelial cells. We performed CCA to identify common sources of variation between datasets and generated a combined Seurat object. To ensure the inclusion of only relevant clusters, we excluded any clusters that did not express marker genes associated with canonical keratins were excluded. This step helped to refine the analysis and focus on clusters that exhibit epithelial characteristics. Finally, we specifically identified ciliated epithelium cells within the dataset using well-established markers such as *FOXJ1*, *PIFO* and *TPPP3*. By leveraging these markers, we conducted relevant analyses to explore the characteristics and functionality of ciliated epithelial cells in the context of endometriosis.

### Cell-cell interaction analysis

We conducted Cell-cell interaction analysis using CellChat^76^ (v.1.6.1). To ensure data integrity, we performed quality inspection and normalization on both the control and adenomyosis datasets, Following the official workflow and default parameter settings, we loaded the control and adenomyosis datasets separately into CellChat. To identify potential ligand-receptor interactions between cell types in the control and adenomyosis datasets, we utilized the built-in CellChatDB.human database as a reference. This database contains known information about receptor-ligand interactions in human cells. By leveraging this reference, we screened for specific interactions and examined their potential significance in the context of control and adenomyosis samples. To quantify the strength and significance of the potential ligand-receptor interactions, we employed various functions within CellChat. The computeCommunProb function was used to calculate the communication probabilities between different cell types. Additionally, we utilized the computeCommunProbPathway function to identify potential pathways associated with the observed interactions. The aggregateNet function was employed to integrate and analyze the resulting network.

### Statistical analysis

Statistical analysis was performed using the R software (v.4.2.2). For all variables, we applied one-tailed wilcoxon rank-sum test to assess the differences between groups. A significance threshold of *P* < 0.05 was used to determine statistical significance. Results that met this criterion were considered to have a statistically significant difference.

## Data Availability

All data can be viewed in NODE (https://www.biosino.org/node)

